# Interpretable machine learning model for data driven classification of Oral Health Related Quality of Life in Patients with Type 2 Diabetes Mellitus

**DOI:** 10.1101/2024.05.03.24306811

**Authors:** Roomani Srivastava, R Murali, Meena Jain, Kshitij Jadhav

## Abstract

Type 2 Diabetes Mellitus(T2DM) is a debilitating condition with a number of complications including those of the oral cavity which can further deteriorate patient’s general and oral health related quality of life (OHRQoL). Machine Learning (ML) can help assign an individual’s propensity to develop poor OHRQoL, given a set of variables, and at the same time identify the most important features contributing to this outcome. Previously inferential statistical methods have attempted to explain this, albeit with limited success. The aim of this cross sectional study is to determine the impact on OHRQoL in T2DM patients, and identify features most likely to be associated with this outcome and to compare ML and DL analytical methods with inferential statistics. Twelve-hundred T2DM patients were subjected to OHRQoL and demographic data questionnaires and WHO Oral Health Assessment form. K-means Clustering was performed to label individuals as having or not having an impact on OHRQoL. Class imbalance was addressed by undersampling of the majority class using informed subset selection. Further, using the collected data as input features we developed ML algorithms (Naive Bayes(NB), Random Forest(RF), Logistic Regression(LR), Kernel Support Vector Machine(SVM) and Artificial Neural Network(ANN)), to accurately classify individuals with or without poor oral health related quality of life (OHRQoL) and utilized SHapley Additive exPlanations (SHAP) analysis for feature importance. The best performing model was SVM (AUC=0.983; Sensitivity=1) for classifying the patients into into poor OHRQoL. SHAP values were highest for Age, Prosthetic Need, Tobacco use and years since onset of diabetes. Features closely related to diabetes, that is, periodontal pockets and loss of attachment were not identified as relevant by inferential statistics, but were deemed as important features associated with poor OHRQoL by SHAP analysis.

## 1 Introduction

Substantial research has established that several systemic disorders have oral manifestations contributing to overall deteriorating health of an individual [1]. Diabetes Mellitus is one such condition which has a well-established bidirectional relationship with oral health [2]. Type 2 Diabetes Mellitus (T2DM) is a chronic debilitating condition that requires continual medical care, ongoing patient self-management, education and support to prevent complications [3] sometimes requiring multifactorial risk reduction strategies beyond glycaemic control [4].

At a prevalence of 75 million, India is considered the diabetes capital of the world with estimated increase to 125 million by 2045 [5]. Of these potentially less than half (45.8 %) are aware of their diabetic status and even fewer (36.1 %) are on treatment [6]. Complications associated with T2DM (cardiovascular, microvascular, renal, ophthalmic) warrant its effective management. A recent review reported that data from High Income Countries (HICs) reflects a reduction in these complications, whereas information on this aspect from Low and Middle Income countries (LMICs) is sparse. However, even limited data reports an increase in complications [7]. Often overlooked complications of diabetes related to the oral cavity include conditions like periodontitis, xerostomia, tooth loss, gingivitis, odontogenic abscess, dental caries and opportunistic infections of tongue and oral mucosa. [2]. Most of these complications have a negative impact on overall health and well-being of an individual.

Health is viewed today in terms of the biopsychosocial dimensions which incorporates factors such as emotional and social well-being along with symptoms and physical measurable health, all of which encompasses Quality of life (QoL) [8], described as the individual’s “perceptions of their position in life in the context of culture and value systems in which they live, and in relation to their goals, expectations, standards, and concerns” [9]. This is now considered an indispensable parameter for assessment of patients’ status in all fields of healthcare, including oral health. Efforts have been invested in developing instruments to measure OHRQoL and Oral Health Impact Profile −14 (OHIP-14) is one such instrument[10] which measures individual’s perception of the social impact of oral disorders on their well-being.

A number of oral health factors have been linked with poor OHRQoL and this has been reported by several studies internationally [11–15] and a few from India [16, 17]. However empirical evidence as to which factors are truly affecting OHRQoL is lacking. An insight into this will allow dentists to focus on specific issues among patients with T2DM which are likely to affect them the most in their day to day functioning. Novel tools such as Machine Learning (ML) and Deep Learning (DL), can be used to achieve this. While previous research has utilized traditional statistics to identify these factors, there has been no investigation that utilizes ML driven modeling to identify factors that are associated with worsened QoL within the cohort of individuals with T2DM. This is critical from an interventional perspective so that early warning triggers can be identified to develop strategies which could alter the course of development of poor OHRQoL in T2DM patients which contributes substantially to the disease burden.

The aim of the present study was to build a Machine Learning classifier to effectively delineate patients whose OHRQoL was impacted negatively from those whose weren’t, given set of oral and sociodemographic features and also determine which of these features are most likely to classify this outcome correctly. A secondary objective was to compare ML and DL analytical methods with inferential statistics.

## 2 Methods

This cross-sectional analytical study was conducted in the outpatient department of Government General Hospital in Bangalore, India. The sampling frame was 35-74 year old patients diagnosed with T2DM for at least the past 3 years who reported to the outpatient department of Government General Hospital Yelahanka, Bangalore. Subjects with known Insulin Dependent Diabetes Mellitus/Gestational diabetes or those with immunosuppressive diseases receiving corticosteroids were excluded from the study.

### 2.1 Sample Size Estimation

The estimated sample size for the proposed study is 1200, which was obtained as per the following formula.

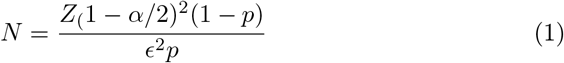

Where, *N* = sample size, *Z* = 1.96 when *α* is assumed to be 0.05, *ϵ* = 0.15, variance estimated to be 15%, *p* = 25%, prevalence of poor OHRQoL among diabetics based on our pilot study done on 50 study subjects. This resulted in a sample size of 1,144, with a drop-out rate of 5%, the final sample size was 1200 individuals.

### 2.2 Ethical Approval and permissions

Ethical clearance was obtained from the Institutional Review Board (Ref No.: 02_D012_44817) of a dental school associated with the general hospital.

### 2.3 Collection of Data

Data collection consisted of interviewing the study subjects regarding their OHRQoL using an instrument OHIP-14 [10] which is based on Locker’s conceptual model of oral health [18]. This was followed by assessing the oral health status assessment using the WHO Oral Health Assessment form 1997 [19]. Further details on data collection are available in the Supplementary Section.

### 2.4 Data Analysis

#### Defining the Clusters

The Total OHIP Score ranges from 0 to 56. Authors of this index have not outlined any scoring criteria for labeling an individual to have good or poor OHRQoL[10]. Most of the studies utilizing OHIP till date have heuristically divided individuals as having an impact or no impact OHRQoL[20]. However, to minimize human bias in determining this threshold, we followed a data driven approach to divide the dataset into these two classes. We used K-means clustering on total OHIP scores [21] to delineate spaces in a multidimensional space in order to assign, each individual (data point) to a cluster which was labeled either as having poor OHRQoL or good OHRQoL.

#### Data Preprocessing

Data collected was first reviewed and only those independent variables were retained as features (driven by domain knowledge) which could have an established relationship with the outcome, OHRQoL, in individuals with T2DM. Data was subjected to preprocessing tools to make it amenable to ML analysis. All categorical variables were one-hot-encoded[22]. Thirty-two input features were thus established. Feature Scaling was performed after splitting the data (test size = 0.3), in order to make all variables of the training set assume the same scale.

Following K means clustering the size of the two clusters obtained were 875 and 291 respectively, that is a highly imbalanced dataset which can lead to erroneous classification in an ML model wherein the model tends to ignore the minority class [23], potentially resulting in poor sensitivity. This is due to the tendency of the algorithms to prioritize achieving good accuracy without paying heed to class wise accuracy. To tackle this we performed under sampling of the majority class by performing informed subset selection using facility location from the Submodlib library [24]. These selected samples and all minority samples were then concatenated to result in a single training dataset with equal number of datapoints in each class. Subset selection was applied only on the training data set and this subset of the data was used as the final dataset for analysis.

#### Data Analysis

We used four ML and one DL model. For ML we used models which use different underlying principles such as Bayesian logic - Naive Bayes(NB); tree based methods - Random Forest(RF); function based models - Logistic Regression(LR) and Kernel Support Vector Machines(Kernel SVM) all from the ScikitLearn Python library[25]. In DL we used Artificial Neural Network model (ANN) from the TensorFlow library of Python[26].

GridSearchCV paired with KFold cross-validation (10 folds) was implemented to optimize hyperparameters across multiple models. For example in SVM, we tested 600 configurations involving parameters such as C, kernel type, and gamma. Similarly, for other models like Random Forest and Logistic Regression, parameters such as tree depth and regularization constants were fine-tuned. For the ANN, we adjusted factors including the number of neurons, L2 regularization, and epochs, implementing early stopping to prevent overfitting. These techniques ensured our models were robust and generalized, effectively addressing overfitting through k-fold cross-validation. We performed this evaluation across ten distinct iterations with shuffled data partitions that is ten train-test splits (70:30) with random seeds. Accuracy, precision, recall (sensitivity), Specificity and F1 score were calculated for each iteration and average values were presented. ML and Dl models were also implemented on the full, imbalanced dataset in order to highlight improved metrics with subset selection.

SHapley Additive ePlanations (SHAP values) [27] were calculated for the best performing model. Mean absolute SHAP values were derived and plotted to present a global explainability of the model.

Inferential statistics was done to determine the utility of ML algorithms over traditional statistics. Mann-Whitney-U test was done to compare continuous variables and Binary logistic regression was done using Backwards Wald’s method to determine the final variables which were significant in contributing to the impact on OHRQoL (Statistical Significance = p *<* 0.05). Choosing this method allowed us to draw parallels with feature importance method (SHAP) used in ML analysis.

## 3 Results

Data collection was done for 1200 participants, however, completely edentulous participants (n=34) were excluded from final analysis as several parameters cannot be recorded in such patients. Refer to Tables 1 & 2 of supplementary section 3.

**Table 1.**
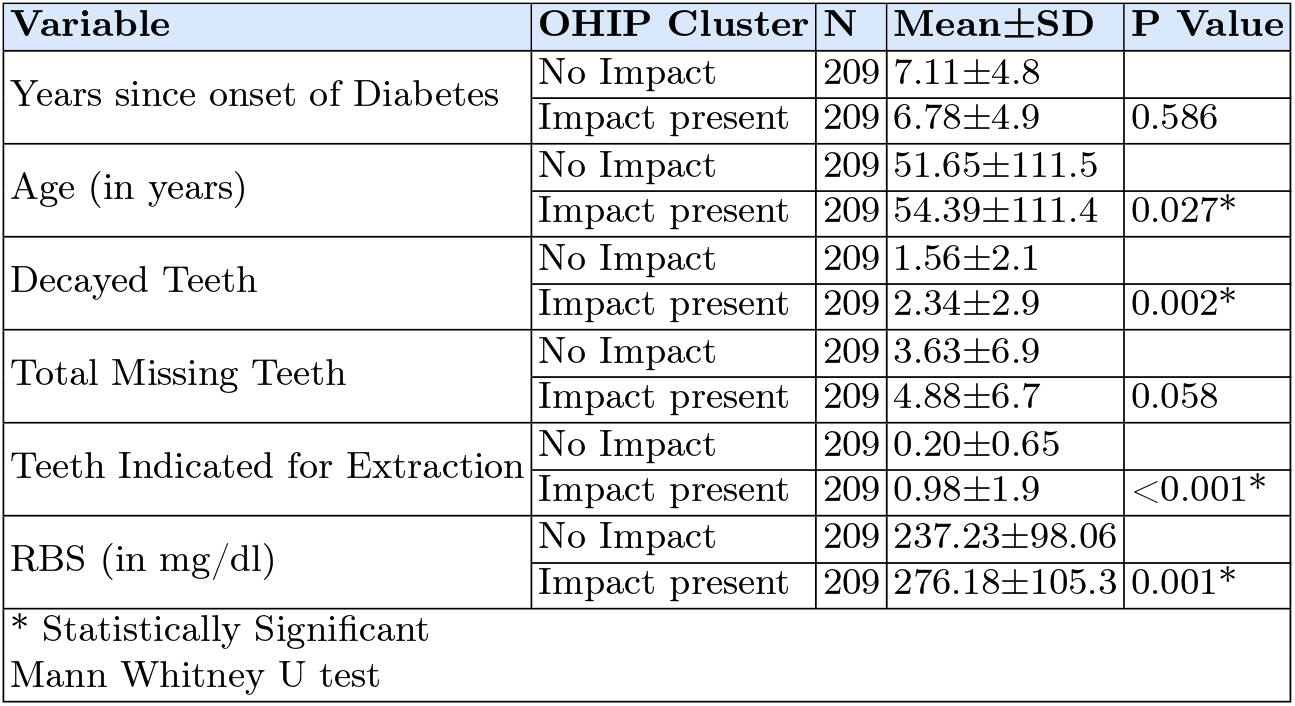
Comparison of continuous variables of the two classes in subset selected data.

**Table 2.**
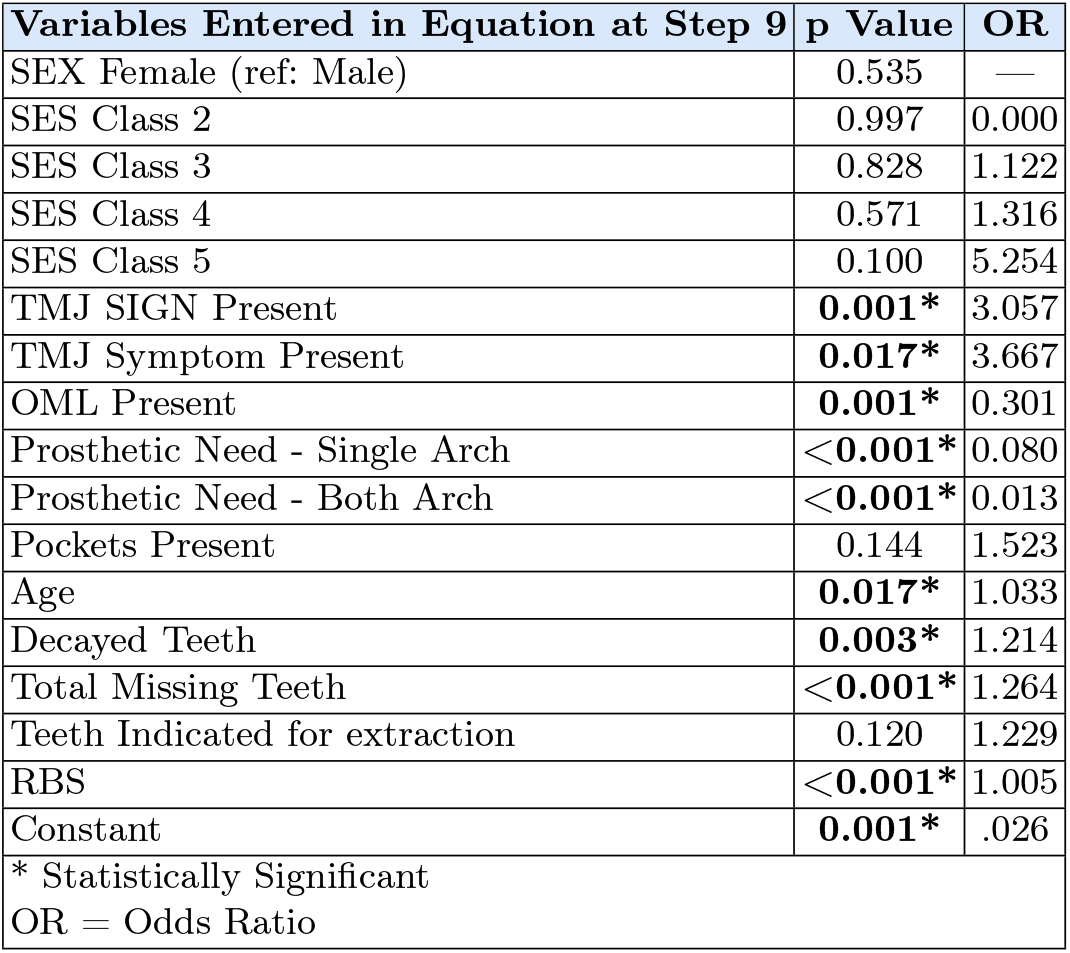
Binary Logistic Regression with OHRQoL as Dependent Variable.

### 3.1 Clustering & Subset Selection

K means Clustering was done to divide datapoints into two clusters. OHIP scores of the **no impact cluster** ranged from 0 to 6.71, with 875 observations (S3 - Table 3). Cluster belonging to **having an impact** on has OHIP scores ranging from 7.06 to 19.2 (n=291). The data was then split into training and test sets in a ratio of 70:30. The training dataset was then subjected to Subset selection by Facility location using the Submodlib library[24] to address the class imbalance. Individuals having impact on OHRQoL (Class 1) in the training dataset were 209; we called this the minority class and those belonging to majority class that i.e. no impact on OHRQoL were 607. The Submodlib algorithm then selected 209 samples from this majority class which best represented it. This subset selected data was then subjected to inferential statistics as well as ML and DL algorithms (figure 1).

**Table 3.**
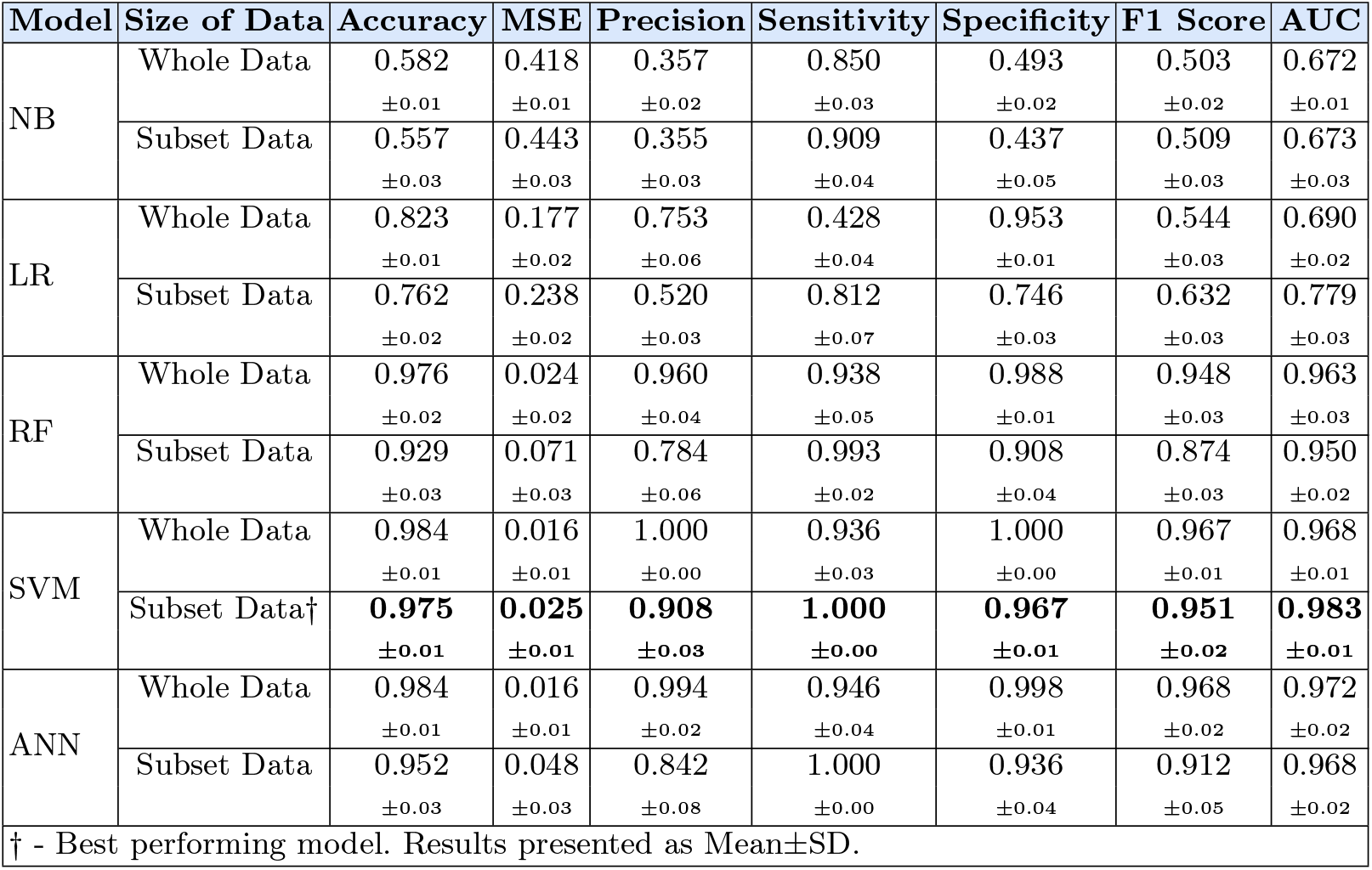
Model Metrics for all the ML & Deep Learning Models.

**Fig. 1.**
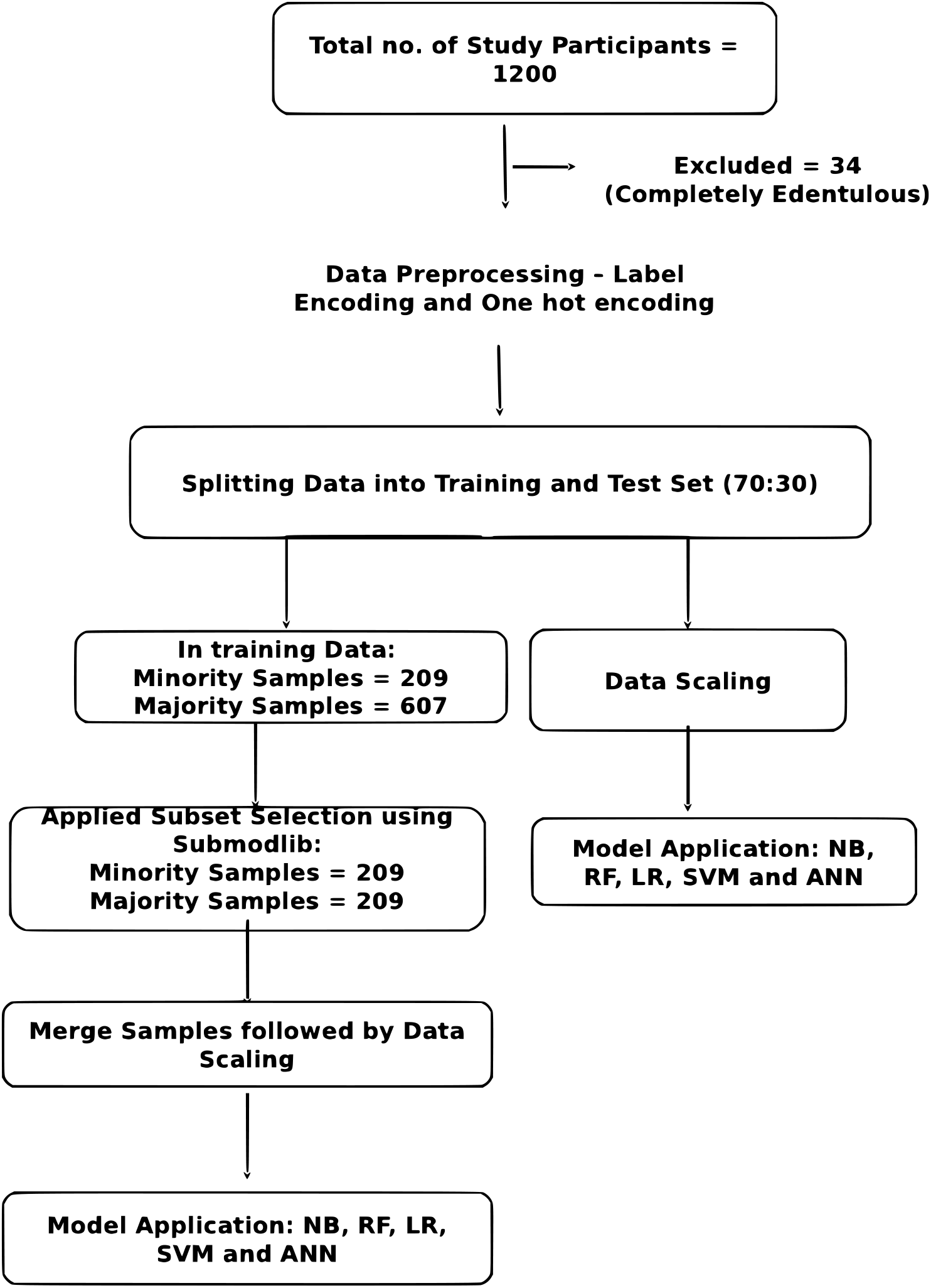
Flowchart of deriving subset of samples and application of algorithms (NB: Naive Bayes, RF: Random Forest, LR: Logistic Regression, SVM: Support Vector Machine, ANN: Artificial Neural Network)

### 3.2 Inferential Statistics Results

Those having an impact on OHRQoL had higher mean age at 54.39 as opposed to those not having an impact which stood at 51.65. The same trend was seen in decayed teeth with ‘Impact’ category having an average of 2.3 decayed teeth v/s 1.5 of no impact category. These differences were statistically significant as per Mann Whitney U test. Teeth indicated for extraction and RBS between the two groups also differed significant, with those having an impact on OHRQoL having RBS values higher by nearly 40 mg/dl. As per Binary logistic regression presence of TMJ signs and symptoms, higher age, more number of decayed & missing teeth and higher RBS all lead to signifcantly higher odds of having an impact on OHRQoL (Table 2).

### 3.3 ML and DL results

AUC improved or remained the same after subset selection for every model other than RF and ANN. However, sensitivity improved after subset selection for each of the models. That is the model’s ability to correctly classify ‘impact on OHRQoL’, improved. SVM-Subset selection was the best performing model with AUC of 0.983 and Sensitivity of 1 (Table 3). The strategy to apply the model after balancing the two classes by subset selection improved the sensitivity of the model, which is rather important in healthcare problems as all diseased conditions must always be classified as diseased even at a cost of a few false positives.

Feature importance was determined using SHAP for the SVM model. They are computed for a specific instance, and can be summed up to provide importance of a single feature in Model performance. SHAP values aim to answer the question: How much did each feature contribute to the difference between the model’s outcome and the expected value of the outcome? A positive SHAP value indicates that the feature has a positive impact on model performance, that is it will lead to correct classification of the condition[27]. Mean absolute SHAP values were positive for all variables(figure 2A) indicating that all input features were essential in classifying an individual as having an Impact on OHRQoL. Features which ranked higher in importance were age, prosthetic need, years since onset of diabetes, decayed teeth, tobacco use, teeth indicated for extraction, TMJ signs & symptoms, low SES and RBS. The beesworm plot (figure 2B) shows all the datapoints in the dataset as a single instance, where blue indicates low feature value and red indicates high feature value. With this it is simple to visualize if data points having positive SHAP value have high or low feature value. For example for the feature ‘age’ most datapoints indicating high value (red) are on the positive side of the plot, that is they have positive SHAP Values, indicating that a higher Age positively impacted the model performance and correctly classified the individual as having an impact on OHRQoL. Periodontal Pockets and Loss of Attachment, two very common conditions amongst diabetic patients were highlighted by the SHAP analysis with positive SHAP values indicating their importance in determining OHRQoL, these factors however were not identified by inferential statistics.

**Fig. 2.**
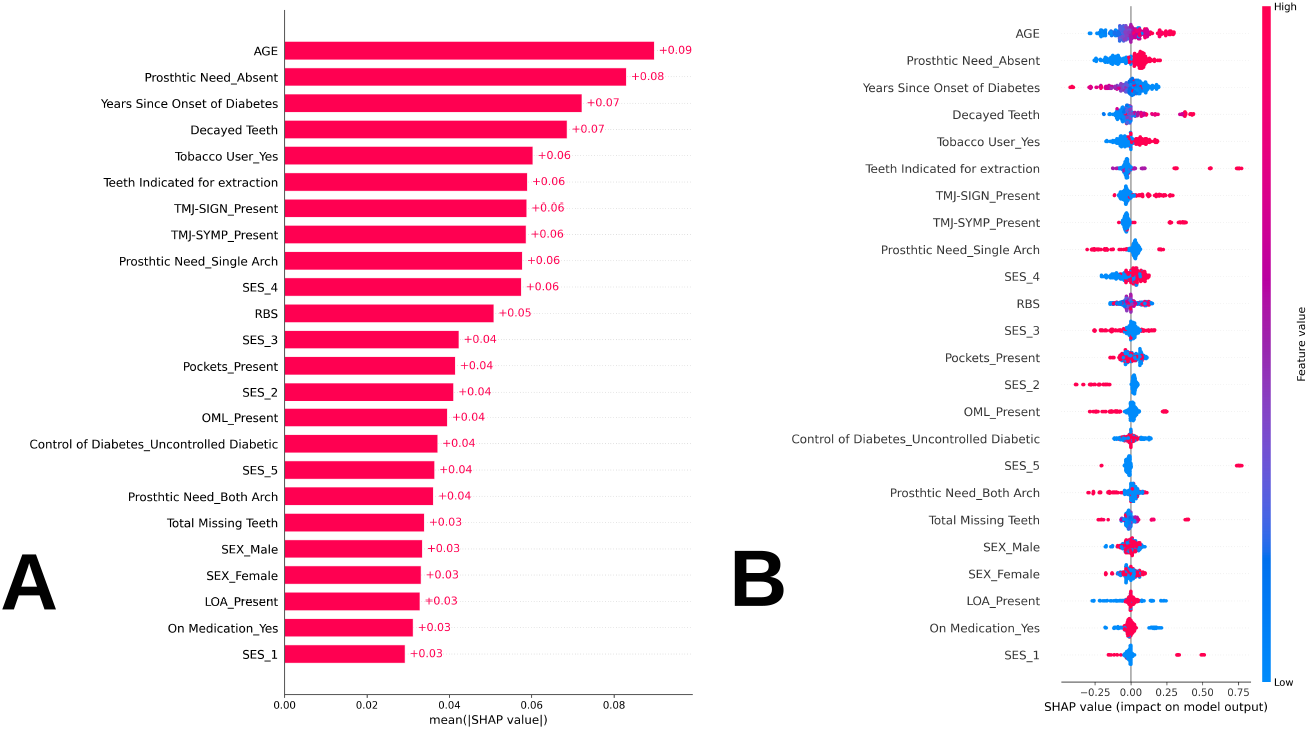
SHapley Additive exPlainations plots: A- SHAP Bar Plot (Values represented are mean absolute SHAP values representing global feature importance); B- SHAP beesworm plot (Individual dots represent the SHAP value of each data point, the colur of the dot denotes weather feature value was high or low)

## 4 Discussion

To the best of our knowledge this is the first study that utilizes the ML based tools to identify which demographic and oral health related factors are strongly associated with poor oral quality of life in individuals with T2DM. In our study, presence of periodontal pockets emerged as a feature for determining OHRQoL in the ML models but not as per inferential statistics. Previous results reported by Sandberg GE et al,[28] were similar to this wherein poor periodontal condition contributed to poor OHRQoL amongst diabetics. However, Pinho et al, [29] reported that difference in prevalence of periodontal disease between groups of people with ‘good’ or ‘poor’ quality of life was not statistically significant. It is prudent to know that research has established a definitive bidirectional relationship between diabetes and periodontal disease[30] and its presence in diabetics is relevant to further course of the disease, eventually affecting OHRQoL.

TMJ signs and symptoms were found to be important factors correctly indicating patients with impact on OHRQoL as per both inferential statistics and ML. The psychological implications of TMJ disorders have been discussed widely in literature along with their bidirectional relationship. Buljan et al [31] have stated that patients with psychiatric problems are 4.5 times more prone to TMJ problems than individuals without psychiatric problems and vice versa. The trigeminal nerve may be affected by diabetic nephropathy in later stages of the disease[32] leading to poor OHRQoL. To the best of our knowledge no other studies have explored the dynamics of OHRQoL and TMJ problems among diabetics. The current study presents an insight worth exploring as early identification of TMJ signs and symptoms could be critical to institute remedial measures to decrease the risk of long term development of neuropsychiatric problems as a secondary outcome of T2DM.

Tobacco use emerged as an important feature classifying patients as having an impact on OHRQoL as per ML but not significant using inferential statistics. Few studies in India have reported the impact of tobacco use and OHRQoL [33, 34] revealing that tobacco use leads to poor OHRQoL. Another study specifically in the diabetic population also reported previous history of tobacco use as an indicator for poor OHRQoL [11]. Tobacco as we know can lead to complications of the microvasculature by itself. A synergism exists between the pathophysiology of diabetes and tobacco use which together can lead to very poor health outcomes in an individual[35]. Additionally, tobacco itself leads to a number of non-cancer oral health outcomes also [36] which include aggravating periodontal disease, attrition leading to tooth sensitivity, all potentially leading to poor OHRQoL. Preemptive action against this combination of diabetes and tobacco use can prevent a myriad of complications in diabetic patients. Both ML and inferential statistical analysis in the present study found that total missing teeth, and teeth indicated for extraction were important in indicating impact on OHRQoL in the study population, also prosthetic need, which is a function of missing teeth, was found to be relevant. This corroborated Busato et al [37] and Ravindranath et al [38] where higher score of DMFT resulted in poor OHRQoL among patients with T2DM. Ravindranath et al also reported being edentulous as a factor. Although, study reported by Sanadhya S et al, 2015 [39] from a different part of the country did not concur as none of the domains showed significant results with respect to prosthetic needs. Having said that, prosthetic need or missing teeth are highly likely to impact day to day living of any individual and more so in diabetics as it affects their diet and nutrition which is essential for the proper maintenance of the disease.

Speaking of the demographic variables a couple of studies [11, 40] also evaluated some variables similar to our study amongst diabetic subjects, where logistic regression analysis revealed that higher age group, lower level of education, poor economic status and duration of diabetes lead to poor OHRQoL. These results seem plausible as Diabetes severity and complications are known to increase with increased age. Moreover variables like education and SES are reflections of level of awareness and access to care[41]. It follows naturally that a combination of these variables will lead to poor OHRQoL.The findings of the above mentioned studies concurred with the present study.

One of the objectives of the present study was to investigate the utility of advanced analytical techniques using machine intelligence as against traditional analytical methods. We identified two major areas where using ML helped over using the standard methods and procedures of inferential statistics. First is in determining the classes of those having an impact on OHRQoL and those not having an Impact on OHRQoL. The instrument used to measure OHRQoL in the present study, the OHIP14, does not have any guidelines for interpreting the scores or any reference ranges to be consulted while classifying patients with good or poor OHRQoL. Studies done utilizing this instrument more often than not heuristically define these categories[20]. In the present study we used a data driven approach to cluster similar values of OHIP. Considering the large sample size, the results of clustering in the present study may be used as a guide by similar studies using the same instrument to group participants into those who had an impact on OHRQoL and those who did not; the cutoff value revealed in this study being 7, thus anyone with a score lower than this can be considered as not having an impact on OHRQoL. Second benefit of using ML methods was observed in terms of determining feature importance. As highlighted above the inferential statistical analysis conducted on the same dataset failed to reveal the relevance of important features such as periodontal pockets, loss of attachment, tobacco use and even gender as contributors to poor OHRQoL. As proven by other research these features do contribute to poor OHRQoL[33, 34, 28] and by using traditional statistical methods, we may not always be able to identify them. Another important aspect to consider is that, inferential statistics utilize the probability principle in rejecting or accepting the null hypothesis[42], so while it may tell us that a feature is important for the outcome of interest, it does not reveal the magnitude of the feature or ranks features in order of importance. SHAP values utilized in ML analysis in this study, on the other hand, reveals a ranking of features based on highest to lowest SHAP value. Another important argument is that most statistical techniques, such as the one used in this study assume linear relationship between variables and fail to perform well when non linear relationships exist. This is captured well by advanced ML and DL models [43]. A further extension of this argument is that feature interactions are also comprehended better by ML analysis. Finally, models developed using ML can be deployed to predict disease, promote health and prevent further regression of disease in real life situations.

The results of the present study can help in identifying a combination of demographic, disease related and oral health related factors which may put an individual at risk of poor OHRQoL which will further deteriorate the already compromised health status of a T2DM patient resulting in aggravating the complications of the disease. Dental needs addressed adequately in such at-risk patients can go a long way in overall disease management. A study done among adults in the United States of America using Structural Equation Modelling [44] analyzed the effects of T2DM, need for dental care, personal health practices and use of services on OHRQoL showing that ‘Dental need’ in T2DM had direct and indirect effects on OHRQoL. This warrants identifying patients at risk of developing poor OHRQoL based on their dental needs, thereby leading to better outcomes in patients with T2DM.

The present study is not without limitations, data collected is from a single region in India. This region was selected owing to the high prevalence of Diabetes in South India. However, data collected from different regions can be collected in the future in order to develop a more generalized model, which could be deployed in practice to delineate patients likely to have poor OHRQoL. Another fact to highlight is that some oral health findings were excluded from analysis as, factors such as enamel hypoplasia, dental trauma, and dental fluorosis, have no documented evidence of their established relationship with presence of diabetes The study aimed to assess only oral complications of diabetes and their effect on OHRQoL.

We used Random Blood Sugar (RBS) test instead of the gold standard HbA1C to ascertain sugar level on the day of examination and as a confirmation of diabetic status whether controlled or uncontrolled. RBS is a useful metric used on day to day basis by those who are already suffering from the condition. It has been postulated that this can be used as a surrogate measure for the control of diabetes, whether poorly controlled or well controlled, as against the gold standard test of HbA1C. More so in resource poor settings such as ours[45].

The cross sectional nature of the study resulting in lack of temporality is an inherent limitation of the study. Longitudinal data on the progress of oral complications of diabetes and OHRQoL can be collected in the future which can provide the opportunity to better capture the progression of the problem and provide more nuanced directions for intervention.

Dental diseases are highly prevalent in India, which results in an excessive overload on the dental health care system and more so in primary care settings where number of consultants are lesser. Control of dental diseases also depends a lot on patient compliance to good oral health practices; this may require behaviour change strategies to be employed by the dentist in counselling and educating such patients. Thus, focusing on at risk patients can lead to a more efficiently running a system for managing oral complications of patients with T2DM.

## Data Availability

All data produced in the present study are available upon reasonable request to the authors, after relevant permissions from parent institution where the study was conducted.

