## Supplementary for "Interpretable machine learning model for data driven classification of Oral Health Related Quality of Life in Patients with Type 2 Diabetes Mellitus"

### Supplementary File

Roomani Srivastava[1[0000−0001−8183−109X]], R Murali[2[0009−0004−5368−9638]], Meena Jain[3[0000−0003−2700−2500]], and and Kshitij Jadhav[1[0000−0001−9795−8335]]

[1] Indian Institute of Technology Bombay, Mumbai, India {22D1628, kshitij.jadhav}@iitb.ac.in
https://www.iitb.ac.in/

[2] Krishnadevaraya College of Dental Sciences and Hospital, Bangalore, India

https://www.kcdsh.org/

[3] Manav Rachna Dental College and Hospital

https://manavrachna.edu.in/mrdc

#### 1 Study settings and data collection

##### 1.1 Study Setting

This cross-sectional analytical study was conducted in the outpatient department of Government General Hospital in Bangalore, India. The sampling frame was 35-74 year old patients diagnosed with T2DM for at least the past 3 years who reported to the outpatient department of Government General Hospital Yelahanka, Bangalore. Subjects with known Insulin Dependent Diabetes Mellitus/Gestational diabetes or those with immunosuppressive diseases receiving corticosteroids were excluded from the study.

A pilot study was conducted with 50 patients reporting to the Hospital which confirmed the feasibility of the methodology of the study. Convenience sampling technique was used to select the study population. Outpatients reporting to the aforementioned hospital having T2DM were examined till the required sample size was achieved.

##### 1.2 Sample Size Estimation

The estimated sample size for the proposed study is 1200, which was obtained as per the following formula.

$$N = \frac{Z_(1-\alpha/2)^2(1-p)}{\epsilon^2 p} \tag{1}$$

Where, $N$ = sample size, $Z = 1.96$ when $\alpha$ is assumed to be 0.05, $\epsilon = 0.15$, variance estimated to be 15%, $p = 25\%$, prevalence of poor OHRQoL among diabetics based on our pilot study done on 50 study subjects (Supplementary Section). This resulted in a sample size of 1,144. A dropout rate of 5% was considered resulting in a final sample size of 1200.

The diabetic status of the patients was confirmed using the medical records of the patients and random blood sugar analysis was done on the day of examination.

Ethical clearance was obtained from the Institutional Review Board (Ref No.: 02_D012_44817) of a dental school associated with the general hospital. The required official permission to select, examine and collect the relevant data from selected subjects had been obtained from the Medical Superintendent of the Government General Hospital. Written informed consent was obtained from each individual before conducting interview and clinical examination.

The principal examiner was trained to ensure uniform interpretation for the various diseases and conditions to be observed and recorded. The examiner practiced the recordings on subjects above 35 years and was calibrated by examining 20 different individuals, and the same examination was repeated after 3-4 hours by the examiner. The results of the two examinations were compared and checked for the intra-examiner reliability (Kappa= 0.88).

##### 1.3 Collection of Data

Data collection consisted of interviewing the study subjects regarding their oral health related quality of life using an instrument OHIP-14 [5] followed by assessing the oral health status assessment using the WHO Oral Health Assessment form 1997 [3]. The assessment form consisted of three sections, the first section consisted of demographic information comprising age, gender, education, income, occupation (for calculation of Socioeconomic Status (SES)) years since onset of diabetes, other medical conditions and the Random Blood Sugar level on that day. The socioeconomic status of the study participants was measured using Kuppuswamy's socioeconomic scale [1]. The Kuppuswamy scale proposed in 1976, measures the SES of an individual based on three variables namely, education and occupation of the head of the household and income of the family. The second section consisted of the Oral Health Impact Profile Questionnaire–14, for the assessment of Oral Health Related Quality of Life. This instrument is based on Locker's conceptual model of oral health [2] which states that any impairment will lead to functional disability resulting in pain and discomfort which may be physical as well as psychological eventually resulting in disability and handicap in the physiological as well as social realm. Thus, based on this conceptual model the OHIP questionnaire has 7 domains namely Functional Limitation, Physical Pain, Psychological Discomfort, Physical Disability, Psychological Disability, Social Disability and Handicap. There are 2 questions in each domain with certain weights assigned to them. Some examples of questions are "Do you have any pain due to problems related to your teeth, mouth or gums" or "Do you have to interrupt meals due to problems with your teeth, mouth and gums". Responses are recorded on a Likert-type scale and coded 4= 'very often', 3 = 'fairly often', 2 = 'occasionally', 1='hardly ever' and 0='never'. Coded responses to each question were multiplied by the weights and the products were added to produce seven subscale scores, the total of which represents the OHIP score.

The third section consisted of WHO Oral Health Assessment Form 1997, which is the most widely used format for oral health assessment. It encompasses all the components of oral health within itself and permits the examiner to have a comprehensive view of the oral health condition of an individual along with the treatment needs. Factors such as number of decayed teeth, presence or absence of TMJ problems, oral mucosal lesions, pockets and loss of attachment, need for prosthesis were collected. Total number of missing teeth were also recorded irrespective of the cause of tooth loss.

Factors such as enamel hypoplasia, dental trauma, and dental fluorosis, though collected, were not used in the final analysis as there is no documented evidence of their established relationship with presence of diabetes. Random Blood Sugar (RBS) test is a useful metric used on day to day basis by those who are already suffering from the condition. It has been postulated that this can be used as a surrogate measure for the control of diabetes, whether poorly controlled or well controlled, as against the gold standard test of HbA1C. More so in resource poor settings such as ours[4]. This RBS was used in the present study as a measure of control of diabetes.

##### 1.4 Limitations of the study methods

The cross sectional nature of the study resulting in lack of temporality is an inherent limitation of the study alongwith lack of non-diabetic control group. The OHIP14 instrument was administered as a guided interview. This method has an inherent limitation that subjects may not be completely frank in their responses. However a study done to assess differences in responses to the OHIP14 used as a questionnaire or in an interview revealed that total OHIP14 scores were not influenced by the method of administration and the use of the OHIP14 in the questionnaire format may result in lower completion rates and loss of data[6].

RBS has been used in this study to assess sugar levels amongst patients with diabetes, for the purposes of simplicity and in order to obtain immediate results. This however may not be as accurate an estimate of blood sugar as compared to the gold standard of glycosylated hemoglobin (HbA1C).

#### 2 Supplementary tables

Table 1: Frequency Distribution of Categorical Variables

| Feature/ Variable | Category | Full Dataset | | Subset Selected Data | |
|---|---|---|---|---|---|
| | | Frequency | Percent | Frequency | Percent |
| Sex | Female | 386 | 33.1 | 160 | 38.3 |
| | Male | 780 | 66.9 | 258 | 61.7 |
| | Total | 1166 | 100 | 418 | 100.0 |
| Socio-Economic Status | Upper Class | 81 | 6.9 | 29 | 6.9 |
| | Upper Middle Class | 139 | 11.9 | 31 | 7.4 |
| | Middle Class | 293 | 25.1 | 102 | 24.4 |
| | Upper Lower Class | 631 | 54.1 | 247 | 59.1 |
| | Lower Class | 22 | 1.9 | 9 | 2.2 |
| | Total | 1166 | 100 | 418 | 100.0 |
| Tobacco Use | No | 706 | 60.5 | 231 | 55.3 |
| | Yes | 460 | 39.5 | 187 | 44.7 |
| | Total | 1166 | 100 | 418 | 100.0 |
| On Medication | No | 189 | 16.2 | 70 | 16.7 |
| | Yes | 977 | 83.8 | 348 | 83.3 |
| | Total | 1166 | 100 | 418 | 100.0 |
| TMJ SIGNS | Absent | 941 | 80.7 | 322 | 77.0 |
| | Present | 225 | 19.3 | 96 | 23.0 |
| | Total | 1166 | 100 | 418 | 100.0 |
| TNJ Symptoms | Absent | 1088 | 93.3 | 383 | 91.6 |
| | Present | 78 | 6.7 | 35 | 8.4 |
| | Total | 1166 | 100 | 418 | 100.0 |
| OML | Absent | 963 | 82.6 | 362 | 86.6 |
| | Present | 203 | 17.4 | 56 | 13.4 |
| | Total | 1166 | 100 | 418 | 100.0 |
| Prosthetic Need | Not Needed | 651 | 55.8 | 252 | 60.3 |
| | Needed in One Arch | 244 | 21 | 70 | 16.7 |
| | Needed in Both Arches | 271 | 23.2 | 96 | 23.0 |
| | Total | 1166 | 100 | 418 | 100.0 |
| Control of Diabetes | Well Controlled | 483 | 41.4 | 166 | 39.7 |
| | Uncontrolled | 683 | 58.6 | 252 | 60.3 |
| | Total | 1166 | 100 | 418 | 100.0 |
| Loss of Attachment | Absent | 249 | 21.4 | 72 | 17.2 |
| | Present | 917 | 78.6 | 346 | 82.8 |
| | Total | 1166 | 100 | 418 | 100.0 |
| Periodontal Pockets | Absent | 504 | 43.2 | 189 | 45.2 |
| | Present | 662 | 56.8 | 229 | 54.8 |
| | Total | 1166 | 100 | 418 | 100.0 |

**Table 2.** Mean and Standard Deviation of Continuous Variables

| Variable | Full Dataset (N = 1166) | Subset Selected Data (N = 418) |
|---|---|---|
| | Mean±SD | Mean±SD |
| Years Since Onset of Diabetes | 6.91±4.7 | 6.9±4.84 |
| Age (in years) | 51.64±11.4 | 53.02±11.5 |
| Decayed Teeth | 1.72±2.2 | 1.95±2.5 |
| Total Missing Teeth | 3.93±6.8 | 4.26±46.8 |
| Teeth Indicated for extraction | 0.37±1.09 | 0.59±1.5 |
| RBS (in mg/dl) | 248.4±98.4 | 256.71±103.5 |

**Table 3.** Clusters defined based on Total OHIP Score using K means clustering

| Category | Minimum | Maximum | N |
|---|---|---|---|
| No impact on OHRQoL | 0 | 6.71 | 875 |
| Impact on OHRQoL | 7.06 | 19.2 | 291 |
| Total | 0 | 19.2 | 1166 |
